# Associations between SARS-CoV-2 variants and risk of COVID-19 hospitalization among confirmed cases in Washington State: a retrospective cohort study

**DOI:** 10.1101/2021.09.29.21264272

**Authors:** Miguel I. Paredes, Stephanie M. Lunn, Michael Famulare, Lauren A. Frisbie, Ian Painter, Roy Burstein, Pavitra Roychoudhury, Hong Xie, Shah A. Mohamed Bakhash, Ricardo Perez, Maria Lukes, Sean Ellis, Saraswathi Sathees, Patrick C. Mathias, Alexander Greninger, Lea M. Starita, Chris D. Frazar, Erica Ryke, Weizhi Zhong, Luis Gamboa, Machiko Threlkeld, Jover Lee, Evan McDermot, Melissa Truong, Deborah A. Nickerson, Daniel L. Bates, Matthew E. Hartman, Eric Haugen, Truong N. Nguyen, Joshua D. Richards, Jacob L. Rodriguez, John A. Stamatoyannopoulos, Eric Thorland, Geoff Melly, Philip E. Dykema, Drew C. MacKellar, Hannah K. Gray, Avi Singh, JohnAric M. Peterson, Denny Russell, Laura Marcela Torres, Scott Lindquist, Trevor Bedford, Krisandra J. Allen, Hanna N. Oltean

**Affiliations:** Department of Epidemiology, University of Washington, Seattle, WA, USA; Vaccine and Infectious Disease Division, Fred Hutchinson Cancer Research Center, Seattle, Washington, USA; Washington State Department of Health, Shoreline, WA USA; Institute for Disease Modeling, Bill and Melinda Gates Foundation, Seattle, WA USA; Department of Laboratory Medicine and Pathology, University of Washington, Seattle, WA, USA; Department of Genome Sciences, University of Washington, Seattle, WA, USA; Brotman Baty Institute for Precision Medicine, Seattle, WA USA; Altius Institute for Biomedical Sciences, Seattle, WA USA; Department of Cardiovascular Services, Swedish Medical Center, Seattle, WA USA; Howard Hughes Medical Institute, Seattle, WA USA

**Keywords:** COVID-19, SARS-CoV-2, Variants, Hospitalization, Vaccination

## Abstract

**Background:** The COVID-19 pandemic is dominated by variant viruses; the resulting impact on disease severity remains unclear. Using a retrospective cohort study, we assessed the hospitalization risk following infection with seven SARS-CoV-2 variants.

**Methods:** Our study includes individuals with positive SARS-CoV-2 RT-PCR in the Washington Disease Reporting System with available viral genome data, from December 1, 2020 to January 14, 2022. The analysis was restricted to cases with specimens collected through sentinel surveillance. Using a Cox proportional hazards model with mixed effects, we estimated hazard ratios (HR) for hospitalization risk following infection with a variant, adjusting for age, sex, calendar week, and vaccination.

**Findings:** 58,848 cases were sequenced through sentinel surveillance, of which 1705 (2.9%) were hospitalized due to COVID-19. Higher hospitalization risk was found for infections with Gamma (HR 3.20, 95%CI 2.40-4.26), Beta (HR 2.85, 95%CI 1.56-5.23), Delta (HR 2.28 95%CI 1.56-3.34) or Alpha (HR 1.64, 95%CI 1.29-2.07) compared to infections with ancestral lineages; Omicron (HR 0.92, 95%CI 0.56-1.52) showed no significant difference in risk. Following Alpha, Gamma, or Delta infection, unvaccinated patients show higher hospitalization risk, while vaccinated patients show no significant difference in risk, both compared to unvaccinated, ancestral lineage cases. Hospitalization risk following Omicron infection is lower with vaccination.

**Conclusion:** Infection with Alpha, Gamma, or Delta results in a higher hospitalization risk, with vaccination attenuating that risk. Our findings support hospital preparedness, vaccination, and genomic surveillance.

**Summary:** Hospitalization risk following infection with SARS-CoV-2 variant remains unclear. We find a higher hospitalization risk in cases infected with Alpha, Beta, Gamma, and Delta, but not Omicron, with vaccination lowering risk. Our findings support hospital preparedness, vaccination, and genomic surveillance.

## Introduction

Following initial detection, SARS-CoV-2 disseminated rapidly worldwide, with the first reported COVID-19 case in the United States detected in Washington State (WA) on January 19, 2020 (1). During the third quarter of 2020, distinct phenotypic changes on the SARS-CoV-2 spike protein were identified, raising concerns about increased transmission or greater disease severity (2). The first detections of these variant viruses in WA occurred on January 23, 2021, when the first two cases of Alpha were found in Snohomish County (3).

Since the initial detection of the first cases of the Alpha variant, multiple SARS-CoV-2 variants have been reported in WA. In March 2021, the Washington State Department of Health (WADOH) partnered with multiple laboratories to establish a sentinel surveillance program to monitor the genomic epidemiology of SARS-CoV-2 (4). Given the replacement of ancestral lineages due to increasingly greater effective reproductive numbers, variant viruses now represent the majority of sequenced cases in WA (4).

The rapid emergence of variant viruses has resulted in numerous studies reporting increased transmissibility (5–8). Previous studies have identified an increased risk of hospitalization for both Alpha and Delta in various regions around the world (9–12). However, these studies compared a single variant lineage to an ancestral lineage or to a small aggregated subset of variant viruses, leaving a dearth of knowledge into how risk of severe disease differs among the various lineages.

To address this gap in knowledge regarding healthcare outcomes following infection with a variant lineage, we designed a retrospective cohort study analyzing epidemiologic and genomic data from WA in order to compare the risk of hospitalization among seven SARS-CoV-2 variants.

## Methods

### Study Design

For this retrospective cohort study, we included cases with SARS-CoV-2 positive RT-PCR results in the Washington Disease Reporting System (WDRS) that contained linking information to corresponding sequences in the GISAID EpiCoV database (13,14) with specimen collection dates between December 1, 2020 and January 14, 2022. Sequence quality was determined using Nextclade version 1.0.1 (https://clades.nextstrain.org/). Lineage was assigned using the Pangolin COVID-19 Lineage Assigner version 3.1.20 (https://pangolin.cog-uk.io/); only cases with an assigned PANGO lineage were included. The primary exposure of interest was SARS-CoV-2 variant, corresponding to all variant viruses that were given a Greek letter variant label by the WHO. These were all assigned a Nextstrain clade making the distinction clear (15). Variants with less than ten hospitalization events were excluded, leaving Alpha, Beta, Gamma, Delta, Iota, Epsilon, and Omicron for analysis as well as ancestral viruses for reference. Vaccination data was collected from the WA IIS repository that is maintained by the Office of Immunizations at WADOH.

Cases without a known age, variant or vaccine manufacturer, cases with multiple lineages identified for the same infection, and cases where the linked viral sequence had >10% sequencing ambiguity, were excluded from the study. For cases with multiple specimens sequenced of the same virus, only the first sequenced specimen was used for analysis. The main analysis was limited to cases with specimens sequenced as part of sentinel surveillance.

### Sentinel Surveillance

As part of an initiative to monitor the genomic epidemiology of SARS-CoV-2, WADOH established a sentinel surveillance program with partner laboratories around the state. Laboratories and the percentage of randomly selected positive specimens they submit for sequencing were designated to optimize representation across WA (16). Only PCR positive samples with a cycle threshold (Ct) of 30 or less are selected for sequencing. In addition to these designated sentinel laboratories, specimens were classified as sentinel surveillance if the sequencing laboratory indicated that they were conducting sequencing on randomly selected specimens. Specimens selected for sequencing as part of outbreak investigations, targeted due to travel history, targeted due to known vaccine breakthrough status, or targeted as part of investigations of S-gene target failures were not considered sentinel surveillance.

### Hospitalizations

The primary outcome of interest was COVID-19 hospitalization. COVID-19 hospitalization is defined as a WA resident with confirmed COVID-19-positive lab who is identified as being hospitalized through hospital records, self-report of hospitalization, or linkage with syndromic surveillance hospitalization records (RHINO). If RHINO hospitalization records differ from the hospital record or self-report, the data is manually reviewed to adjudicate. Cases known to be hospitalized for a condition other than COVID-19 (e.g. labor and delivery) are not counted. In addition to the above data curation by WADOH, we additionally exclude cases where a positive viral collection date is more than 14 days after hospitalization in order to prevent misclassification of hospitalizations not attributable to COVID-19. Cases with a record of hospitalization but without an admission date were excluded from the study.

### Covariates

We identified *a priori* confounders that were suspected to be associated with both risk of hospitalization following a COVID-19 infection and the epidemiological risk of acquiring a variant. These included age at sampling (categorized into 10-year increments), calendar week of collection, sex assigned at birth, and vaccination. Vaccination status was made into a three tier variable of 1) “Unvaccinated to <21 days post dose one”, 2) “ ≥21 days post dose one to <21 days post booster”‘, 3) “≥21 days post booster” due to a low number of hospitalized cases having a record of vaccination. We consider active vaccination only after 21 days due to CDC guidance regarding active protection from symptomatic infection only after 14 days (17) and then allowing for an additional 7 days to allow for the development of protection from hospitalization, given that the mean time from symptom onset to hospitalization was found to be about 7 days (18). Our vaccination covariate includes cases with a history of vaccination with BNT162b2, mRNA-1273, and Ad26.COV2. Additionally, cases with a repeat positive test (defined as a case where the specimen collection date was more than 21 days after the first positive test date) were also excluded from the study to reduce confounding from previous immunity.

### Statistical Analysis

We used descriptive statistics to explore characteristics of our sample stratified by SARS-CoV-2 lineage. For all descriptive analyses, we summarized categorical variables as frequencies and percentages.

We estimated the associations between SARS-CoV-2 variants and the risk of COVID-19 hospitalization by calculating hazard ratios (HRs) for the time to hospital admission through a Cox proportional hazard model with mixed effects using ancestral lineages as the reference group. We adjusted the HRs for the covariates of age, sex assigned at birth, calendar week (continuous), and vaccination status. Sex and vaccination status were added as random effects to regularize adjustments for under-represented categories. A likelihood ratio test was used to examine the global effect of variant lineages on hospitalization risk.

In a secondary analysis to analyze how vaccination affected the risk of hospitalization by variant lineage, an interaction term of vaccination*lineage was introduced into the model and reran for those variants found to have the largest sample size and effect magnitude: Alpha, Gamma, Delta, and Omicron. Stratified risk of hospitalization by vaccination status was conditioned on the “Unvaccinated to <21 days post dose one” group for cases infected with an ancestral lineage.

The above analysis was repeated with a subset of the data only including cases infected with Delta (as the reference) or Omicron with a collection date after September 1st, 2021.

In order to account for differences in both model selection and case inclusion, sensitivity analyses were performed using a Cox proportional hazard model with fixed effects and a Poisson regression model for both the subsetted sentinel surveillance-only dataset as well as for the entire case dataset found in WDRS for the same study period. Statistical analyses were performed using R version 3.6.2 (R Project for Statistical Computing).

Analytic code can be found at https://github.com/blab/ncov-wa-variant-severity

## Results

The COVID-19 epidemic in WA shows a distinct trend in the lineage distribution over time (Fig. 1). Early on, the epidemic was predominantly characterized by ancestral lineages, while by March 2021, SARS-CoV-2 variants gained predominance over ancestral lineages.

**Figure 1:**
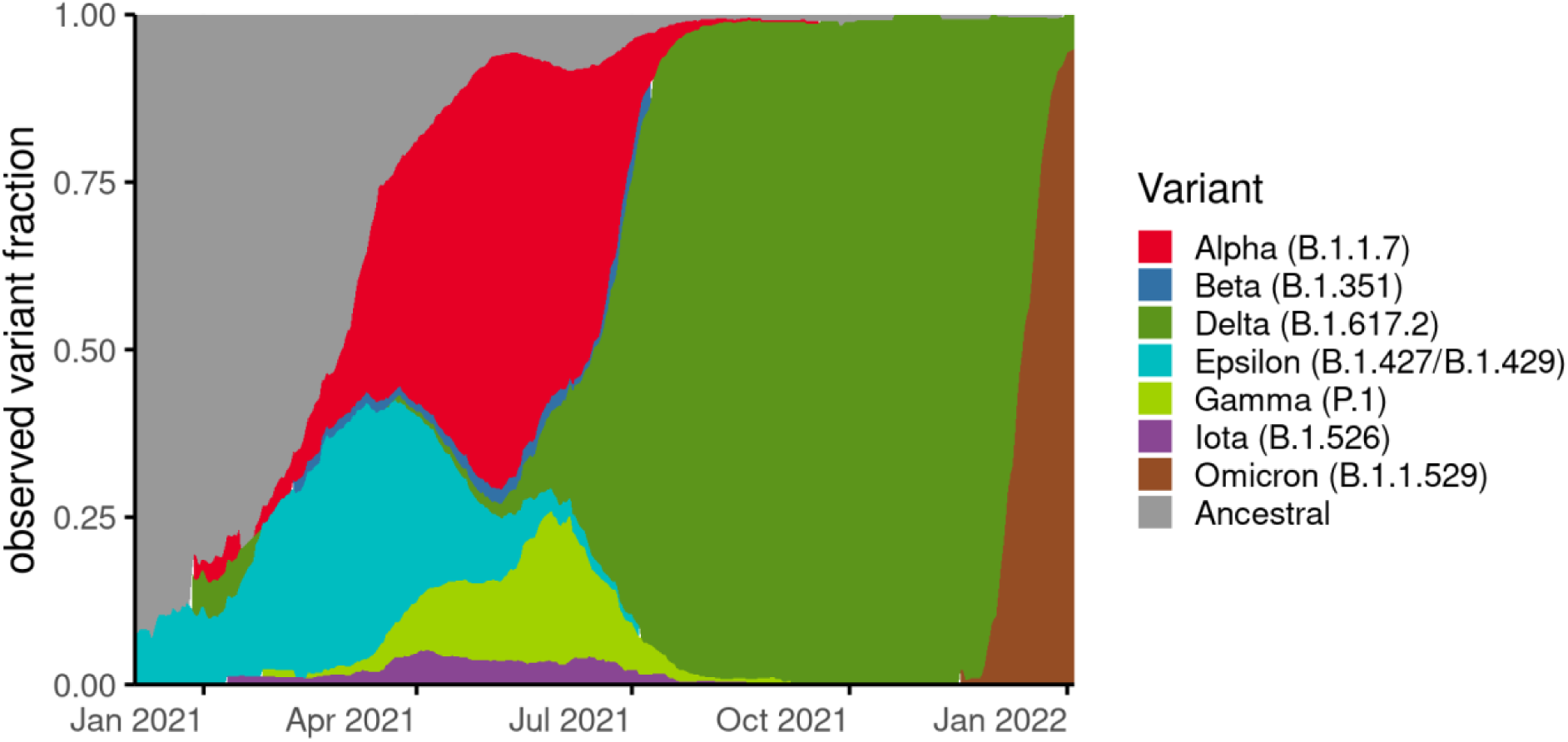
Changing proportion of infections due to variant lineages in Washington over study period. Variant fraction is calculated from a 21-day rolling average from our full sequenced dataset spanning from December 1, 2020 to January 14, 2022 and normalized to 100% to better observe changes in proportion of infections from variant lineages compared to total infections.

In this study, we included 63,639 cases with viral genome data available on WDRS, with specimens collected from December 1, 2020 to January 14, 2022 (Fig. S1). Of these, the final study population for the main analysis was restricted to 58,848 (92.3%) cases that were part of sentinel surveillance. The proportion of total cases in WA that were sequenced as part of sentinel surveillance over time is shown in Fig. S2.

Table 1 represents the general characteristics of the study population. The number of cases infected with a variant includes 8723 (14.8%) infected with Alpha, 231 (0.4%) with Beta, 2101 (3.6%) with Gamma, 33,107 (56.3%) with Delta, and 5362 (9.1%) with Omicron. 5178 (8.8%) individuals were infected with an ancestral lineage other than a variant of concern or interest as defined herein. Of the cases in the main analytic sample, 1705 (2.9%) cases were hospitalized.

**Table 1.** Study Cohort Characteristics by Variant of Concern/Variant of Interest.

| Characteristics | All | Ancestral |  | Alpha |  | Beta |  | Gamma |  | Delta |  | Epsilon |  | Iota |  | Omicron |  |
| --- | --- | --- | --- | --- | --- | --- | --- | --- | --- | --- | --- | --- | --- | --- | --- | --- | --- |
| **Total N\*** | 58,848 | 5178 | (8.8) | 8729 | (14.8) | 231 | (0.4) | 2103 | (3.6) | 33,115 | (56.2) | 3526 | (6.0) | 638 | (1.1) | 5362 | (9.1) |
| **Hospitalized (%)†** | 1705 | 117 | (2.3) | 233 | (2.7) | 11 | (4.8) | 112 | (5.3) | 1109 | (3.3) | 74 | (2.1) | 13 | (2.0) | 36 | (0.7) |
| **Median time (in days) to hospitalization (IQR)** |  | 6 | (5) | 6 | (5) | 7.5 | (3.25) | 6 | (4.75) | 6 | (4.75) | 5 | (4) | 3.5 | (4) | 7 | (6) |
| **Age (%)†** |  |  |  |  |  |  |  |  |  |  |  |  |  |  |  |  |  |
| 0-9 | 5661 | 418 | (8.1) | 888 | (10.2) | 22 | (9.5) | 198 | (9.4) | 3412 | (10.3) | 307 | (8.7) | 54 | (8.5) | 362 | (6.8) |
| 10 -19 | 8954 | 814 | (15.7) | 1561 | (17.9) | 52 | (22.5) | 272 | (12.9) | 4548 | (13.7) | 590 | (16.7) | 113 | (17.7) | 1004 | (18.7) |
| 20- 29 | 12340 | 1053 | (20.3) | 1948 | (22.3) | 51 | (22.1) | 527 | (25.1) | 6485 | (19.6) | 760 | (21.6) | 154 | (24.1) | 1362 | (25.4) |
| 30 - 39 | 11156 | 984 | (19.0) | 1640 | (18.8) | 39 | (16.9) | 432 | (20.5) | 6285 | (19.0) | 657 | (18.6) | 128 | (20.1) | 991 | (18.5) |
| 40 - 49 | 8345 | 741 | (14.3) | 1273 | (14.6) | 34 | (14.7) | 317 | (15.1) | 4583 | (13.8) | 524 | (14.9) | 93 | (14.6) | 780 | (14.5) |
| 50 - 59 | 6012 | 569 | (11.0) | 797 | (9.1) | 22 | (9.5) | 186 | (8.8) | 3529 | (10.7) | 383 | (10.9) | 56 | (8.8) | 470 | (8.8) |
| 60 - 69 | 3807 | 378 | (7.3) | 389 | (4.5) | 9 | (3.9) | 108 | (5.1) | 2429 | (7.3) | 209 | (5.9) | 25 | (3.9) | 260 | (4.8) |
| 70 - 79 | 1704 | 139 | (2.7) | 150 | (1.7) | 2 | (0.9) | 33 | (1.6) | 1223 | (3.7) | 62 | (1.8) | 11 | (1.7) | 84 | (1.6) |
| 80-89 | 681 | 60 | (1.2) | 68 | (0.8) | 0 | (0.0) | 22 | (1.0) | 462 | (1.4) | 31 | (0.9) | 3 | (0.5) | 35 | (0.7) |
| 90+ | 222 | 22 | (0.4) | 15 | (0.2) | 0 | (0.0) | 8 | (0.4) | 159 | (0.5) | 3 | (0.1) | 1 | (0.2) | 14 | (0.3) |
| **Sex (%)†** |  |  |  |  |  |  |  |  |  |  |  |  |  |  |  |  |  |
| Female | 28518 | 2410 | (46.5) | 4234 | (48.5) | 104 | (45.0) | 1026 | (48.8) | 16053 | (48.5) | 1666 | (47.2) | 292 | (45.8) | 2733 | (51.0) |
| Male | 29219 | 2595 | (50.1) | 4328 | (49.6) | 123 | (53.2) | 1040 | (49.5) | 16513 | (49.9) | 1739 | (49.3) | 330 | (51.7) | 2551 | (47.6) |
| Other | 58 | 5 | (0.1) | 9 | (0.1) | 0 | (0.0) | 1 | (0.0) | 36 | (0.1) | 2 | (0.1) | 0 | (0.0) | 5 | (0.1) |
| Unknown | 1087 | 168 | (3.2) | 158 | (1.8) | 4 | (1.7) | 36 | (1.7) | 513 | (1.5) | 119 | (3.4) | 16 | (2.5) | 73 | (1.4) |
| **Vaccination (%)†** |  |  |  |  |  |  |  |  |  |  |  |  |  |  |  |  |  |
| No Vaccination to <21 post dose one | 44845 | 5100 | (98.5) | 8412 | (96.4) | 221 | (95.7) | 1946 | (92.5) | 23108 | (69.8) | 3438 | (97.5) | 620 | (97.2) | 2000 | (37.3) |
| ≥21 days post dose one to <21 days post booster | 12895 | 75 | (1.4) | 309 | (3.5) | 10 | (4.3) | 154 | (7.3) | 9434 | (28.5) | 86 | (2.4) | 18 | (2.8) | 2809 | (52.4) |
| ≥21 days post dose one | 1142 | 0 | (0.0) | 2 | (0.1) | 0 | (0.0) | 1 | (0.1) | 565 | (1.7) | 2 | (0.1) | 0 | (0.0) | 553 | (10.3) |

\*denotes row percentages †denotes column percentages

In the adjusted model, we find a significant global effect of variant lineages on the hospitalization risk when compared to those cases infected with an ancestral virus (likelihood ratio test, p <0.001). The highest risks (Fig. 2) were found in cases infected with Gamma (HR 3.20, 95% CI 2.40-4.26) or Beta (HR 2.85, 95% CI 1.56-5.23). Cases with infection by Delta (HR 2.28, 95% CI 1.56-3.34) or Alpha (HR 1.64, 95% CI 1.29-2.07) also showed a higher risk of hospitalization when compared to the reference. All other variants, including Omicron (HR 0.92, 95% CI 0.56-1.52) failed to show a significant difference in risk of hospitalization (Table 2).

**Figure 2:**
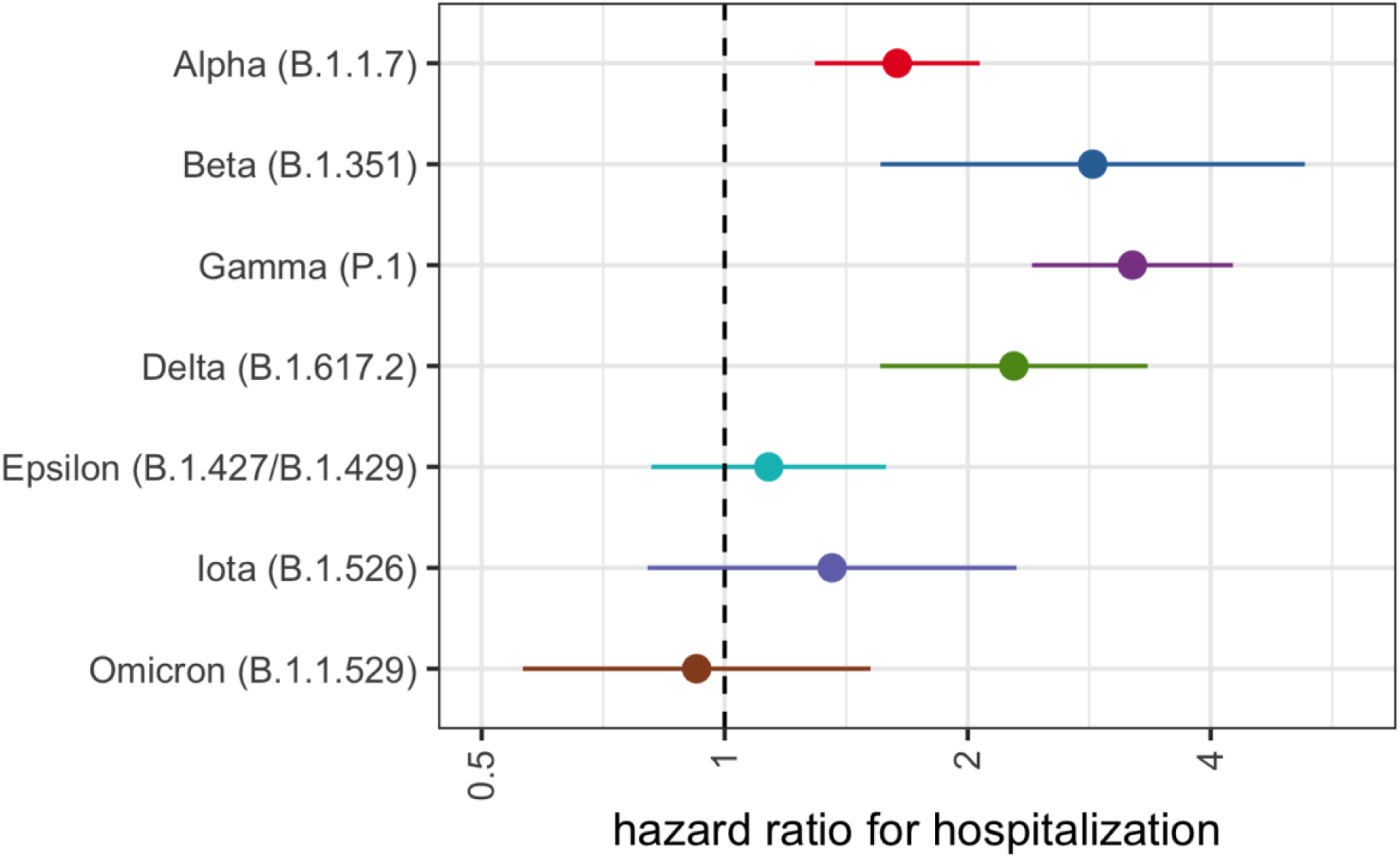
Relative Risk of Hospitalization by Variant Lineage. Risk of hospitalization is compared to individuals infected with an ancestral lineage. Error bars represent 95% CI. Estimates are adjusted for age, sex assigned at birth, calendar week, and vaccination status.

**Table 2:** Adjusted Cox Proportional Hazard Estimates for Risk of Hospitalization.

| Characteristics | Hospitalization |  |
| --- | --- | --- |
|  | HR | 95% CI |
| *WHO Lineage* |  |  |
| Ancestral | REF |  |
| Alpha | 1.64 | (1.29-2.07) |
| Beta | 2.85 | (1.56-5.23) |
| Gamma | 3.20 | (2.40-4.26) |
| Delta | 2.28 | (1.56-3.34) |
| Epsilon | 1.13 | (0.67-1.90) |
| Iota | 1.34 | (0.80-2.30) |
| Omicron | 0.92 | (0.56-1.52) |
| *Vaccination* |  |  |
| Unvaccinated to<br><21 days post dose<br>one | REF |  |
| ≥21 days post dose<br>one to <21 days<br>post booster | 0.40 | (0.35-0.45) |
| ≥21 days post<br>booster | 0.31 | (0.19-0.51) |

\*Additional model covariates include: sex, age (in 10 year bins), calendar week.

The association between variant lineage and hospitalization risk stratified by vaccination is shown in Figure 3 with unvaccinated individuals (unvaccinated or <21 days post dose one) infected with ancestral lineages as the reference category. When compared to the reference, our model shows a higher risk of hospitalization for those unvaccinated individuals infected with Gamma, Delta, or Alpha, while those infected with Omicron showed no significant difference (Table 3). In the strata of individuals with an active vaccination but no active booster, no significant difference was observed in the risk of hospital admittance following infection with Gamma, Delta, or Alpha, but a lower risk of hospitalization was found following infection with Omicron (HR 0.49 95% CI 0.29-0.83), all when compared to unvaccinated, ancestral lineage cases. For those variant categories who had at least 4 hospitalizations following active booster vaccination (Table S1), we find a significantly lower risk of hospitalization for cases infected with Omicron (HR 0.44 95% CI 0.21-0.93) but no significant difference for those infected with Delta, both compared to the unvaccinated, ancestral reference. Without stratification by variant lineage, we find that when compared to the unvaccinated group, cases with a record of an active vaccination but no booster and those with an active booster vaccination both have a lower risk of hospitalization (≥21 days post dose one but <21 days post booster: HR 0.34, 95% CI 0.23–0.50; ≥21 days post booster: HR 0.31 95% CI 0.19-0.51).

**Figure 3:**
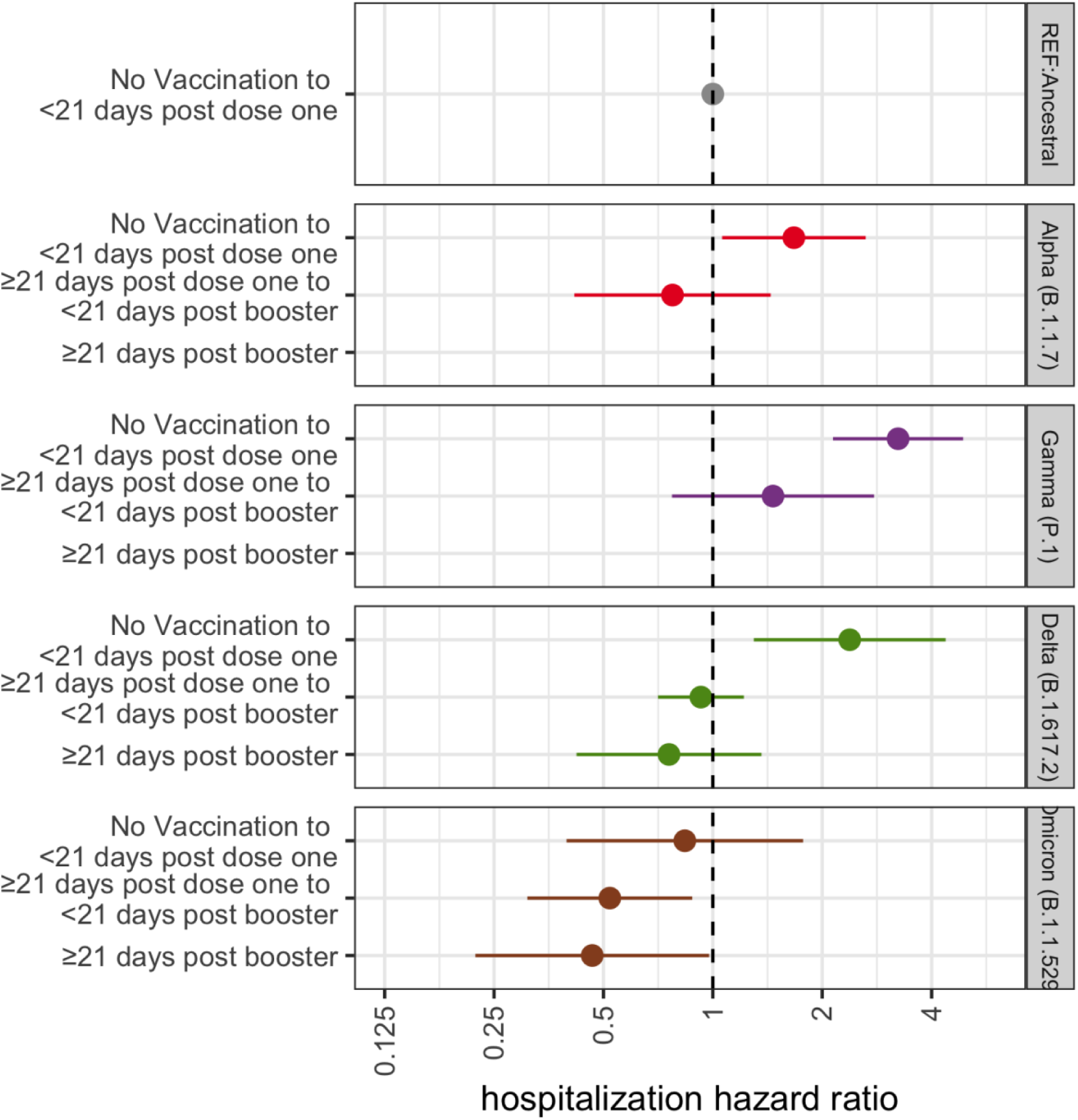
HR for risk of hospitalization following infection with a VOC (excluding Beta due to small sample size) stratified by vaccination status. Unvaccinated individuals infected with ancestral lineages serve as the reference category for each VOC HR. Error bars represent 95% CI. Estimates are adjusted for calendar week, age and sex assigned at birth. Categories with less than 4 hospitalizations are censored.

**Table 3:** Adjusted Cox Proportional Hazards Estimates for Risk of Hospitalization for Variant-Vaccine Interaction.

| Characteristics | Hospitalization |  |
| --- | --- | --- |
|  | HR | 95% CI |
| Vaccine*Variant |  |  |
| *Alpha* |  |  |
| Unvaccinated to <21 days post dose one | 1.67 | (0.91- 3.07) |
| ≥21 days post dose one to <21 days post booster | 0.78 | (0.42-1.42) |
| ≥21 days post booster | — | — |
| *Gamma* |  |  |
| Unvaccinated to <21 days post dose one | 3.24 | (2.15 -4.89) |
| ≥21 days post dose one to <21 days post booster | 1.46 | (0.77-2.78) |
| ≥21 days post booster | — | — |
| *Delta* |  |  |
| Unvaccinated to <21 days post dose one | 2.39 | (1.32-4.32) |
| ≥21 days post dose one to <21 days post booster | 0.93 | (0.71-1.22) |
| ≥21 days post booster | 0.75 | (0.41-1.34) |
| *Omicron* |  |  |
| Unvaccinated to <21 days post dose one | 0.79 | (0.37-1.67) |
| ≥21 days post dose one to <21 days post booster | 0.49 | (0.29-0.83) |
| ≥21 days post booster | 0.44 | (0.21-0.93) |
| *Ancestral* |  |  |
| Unvaccinated to <21 days post dose one | REF |  |

\*Additional model covariates include: sex, age (in 10 year bins), calendar. Each variant lineage category risk estimate uses the “Unvaccinated to <21 days post dose one” vaccination group in cases infected with an ancestral lineage as the reference group. Categories with less than 4 hospitalizations are censored.

In a secondary analysis comparing hospitalization risk following infection with Omicron to infection with Delta as the reference (Fig 4, Table S2), we find a lower risk of hospitalization associated with Omicron infection (HR 0.34, 95% CI 0.23-0.50). When stratified by vaccination status, we find progressively lower risks of hospitalization for cases infected with Omicron for those unvaccinated (HR 0.37, 95% CI 0.21-0.66), vaccinated without a booster (HR 0.23, 95% CI 0.14-0.39), and those ≥21 days post booster (HR 0.19, 95% CI 0.09-0.41), all when compared to unvaccinated cases with Delta infections.

**Figure 4:**
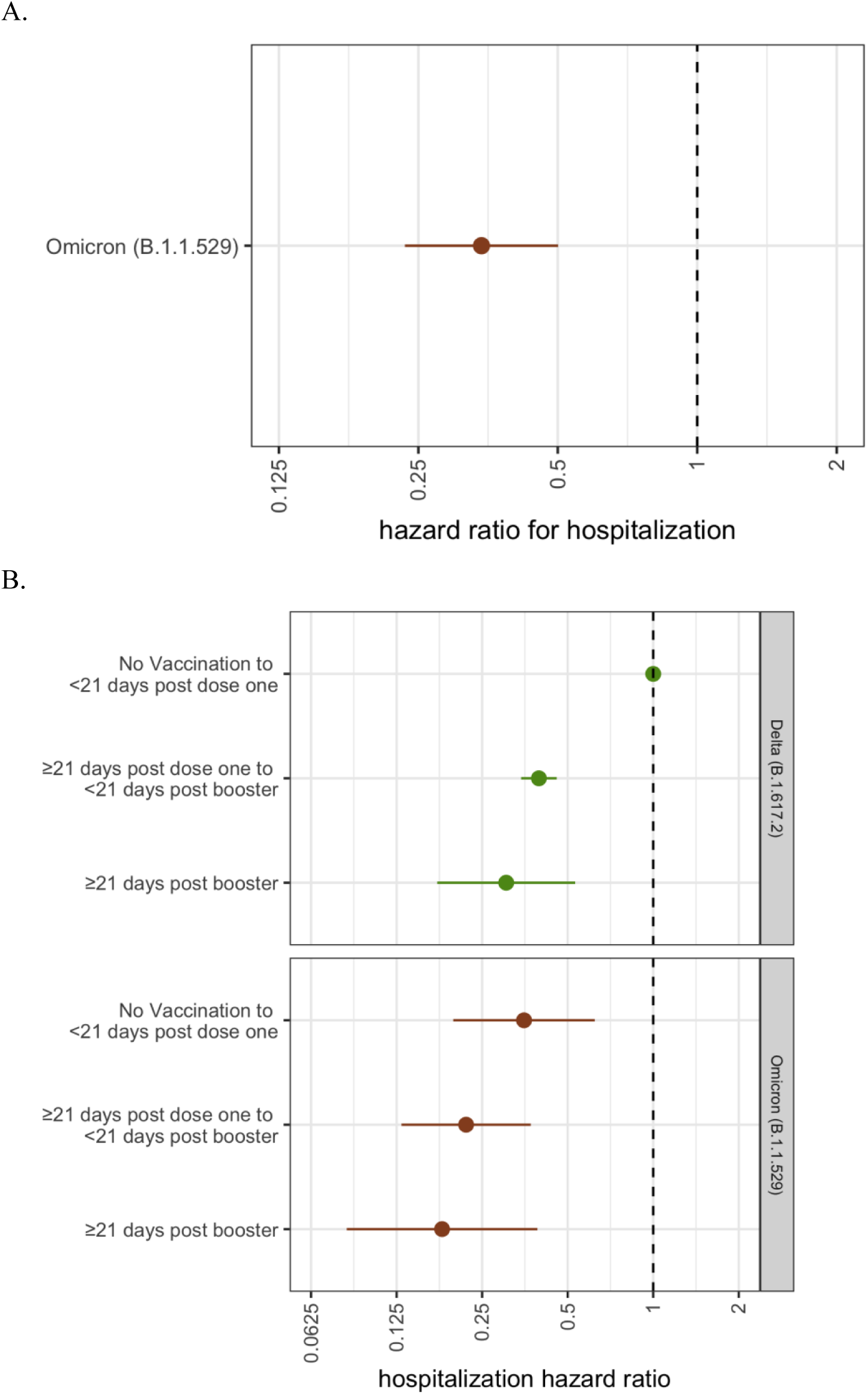
Risk of Hospitalization following Infection with Omicron vs Delta. A. Risk of hospitalization is compared to individuals infected with Delta. B. Unvaccinated individuals infected with Delta serve as the reference category for each VOC HR. Error bars represent 95% CI. Estimates are adjusted for calendar week, age and sex assigned at birth.

Estimates of the HR of the risk of hospitalization for cases infected with variants are robust to both model selection and inclusion of all sequences in our original database (Supp. Fig. 2-3).

## Discussion

In this study, we use SARS-CoV-2 cases in WA that were sequenced as part of sentinel surveillance to evaluate the differential risk of hospitalization following infection with a variant. We find that in our study period, cases infected with Alpha, Beta, Gamma, or Delta have a higher hospitalization risk compared to cases infected with an ancestral lineage, after adjusting for relevant covariates. We find similar estimates of higher hospitalization risk in the subset of unvaccinated individuals and no significant difference in hospitalization risk in individuals with an active vaccination, following infection with Gamma, Delta, or Alpha, while individuals infected with Omicron show a lower hospitalization risk in all vaccination categories, all compared to unvaccinated individuals infected with ancestral lineages.

Our findings are consistent with studies from around the world that have examined hospitalization risk following infection with SARS-CoV-2 variants (11). Our estimates of hospitalization risk following infection with Delta (HR 2.28 95% CI 1.56-3.34) are similar to those from Scotland (HR 1.85 95% CI 1.39-2.47)(12) and Public Health England (HR 2.61, 95% CI 1.56-4.36) (19). To our knowledge, few studies outside of ours have examined the hospitalization risk of Omicron compared to infection with ancestral lineages, but our estimates of hospitalization risk of Omicron vs Delta are highly similar to those calculated using S-gene target failure (SGTF) data (20–22). Unlike studies using only SGTF data to identify probable Omicron cases, our study uses genomic sequencing to confirm the variant identity of each case, reducing the risk of misclassification. Verification with genomic sequencing is crucial for estimating the severity of Omicron, especially given the global rise of the BA.2 sublineage which does not cause S-gene dropout in TaqPath assay.

We also evaluated hospitalization risk following infection by Alpha, Gamma, Delta, or Omicron stratified by vaccination status. Following infection with Alpha, Gamma, or Delta, we saw a higher hospitalization risk for unvaccinated individuals and no significant difference in risk for vaccinated individuals without a booster when compared to those unvaccinated individuals infected with an ancestral lineage. Vaccinated individuals without a booster infected with any of these three variants all showed similar estimates of hospitalization risk with overlapping confidence intervals. The similar, overlapping estimates of risk in vaccinated individuals following infection with a VOC is supported by studies in the United Kingdom and Denmark showing no significant difference in hospitalization risk for vaccinated individuals infected with Delta when compared to those infected with Alpha, together suggesting that vaccination exerts a similar effect across these three variants (11,25). Unvaccinated individuals infected with Omicron showed no significant difference in risk of hospitalization, but individuals with any vaccination were found to have a lower hospitalization risk, all when compared to the unvaccinated, ancestral reference. When comparing hospitalization risk of Omicron vs Delta stratified by vaccination status, we find that any active vaccination is associated with a lower risk of hospitalization regardless of lineage when compared to unvaccinated cases with Delta infections, with estimates of risk similarly observed in Denmark (21).

Our sample sizes in some stratum are small (Table S1) limiting our ability to make conclusions. Additionally, cases were selected into our study based on test positivity; if vaccinated individuals are less likely to seek testing and severe illness leads to increased testing, conditioning study enrollment on testing can lead to collider stratification bias, or a distorted association between vaccination and disease severity (26). Prior to July 27, 2021, CDC guidance stated that fully vaccinated individuals without symptoms did not need to get laboratory tested for SARS-CoV-2 following an exposure, meaning that cases in our sample with a vaccination record, which are conditional on being tested, are almost certainly biased towards a subset of the population with a more severe clinical presentation than the population at-large, potentially underestimating estimates of vaccine protection on hospitalization risk. However, Delta and Omicron estimates largely derive from cases and hospitalizations after July 27, 2021.

Although our findings are consistent with previous studies, they are not without limitations. Variant classification is conditional on whole genome sequencing and a Ct threshold <30, meaning that our sequenced cohort may have been different from the general population of cases in WA. Sample sizes were determined by variant-specific circulation in WA and thus some variant categories have as few as 11 hospitalizations (Table 1: Beta); these respective estimates should be interpreted accordingly. Vaccination data in IIS is not comprehensive of federal vaccination efforts; vaccination status may therefore be misclassified for some cases. Sentinel specimens included in this study were randomly selected for sequencing within laboratories, but laboratories were not randomly sampled for inclusion in the sentinel surveillance program. The implementation of this program set proportions for sampling from specific laboratories to gather a geographically representative sample. It is possible that laboratory-level association with patient populations with differential risk of hospitalization over time may bias the study findings. The study is observational in nature, meaning that despite adjusting for potential confounders, there might be other confounders such as use of monoclonal therapy, social deprivation, etc., that might affect the association between SARS-CoV-2 variant and hospitalization risk. While previous studies have included comorbid conditions, race/ethnicity, or region of residence, their association with the risk of infection with a variant vs an ancestral strain in WA is unclear and thus were not included in our *a priori* set of confounders. Including these variables in an exploratory model did not affect estimates (Delta adjusting for race and county: HR 2.21 95% CI 1.50-3.30; Delta without race and county: HR 2.28 95% CI 1.56-3.34).

In conclusion, our retrospective cohort study found a higher hospitalization risk in cases infected with Alpha, Beta, Gamma, and Delta, but not Omicron. Our study supports hospital preparedness in areas with uncontrolled viral spread as well as promoting vaccination. This study also highlights the importance of ongoing genomic surveillance at the state and federal level to monitor variant outcomes. Building a robust public health workforce as well as collaborations between public health and academia is critical to using genomic epidemiology to answer crucial questions about emerging SARS-CoV-2 variants.

## Data Availability

Requests to access these data are handled by the Washington Department of Health

## Acknowledgments

Clinical and sentinel laboratories who forwarded specimens for sequencing, and sequencing laboratories that reported data to WADOH. The WADOH Data Science Support Unit for integrating sequencing data with epidemiologic case data. The WADOH IIS team and DIQA teams for maintaining and linking vaccination data. The WADOH surveillance team for case and hospitalization data. Natasha Close and the WADOH RHINO team for hospitalization data. James S. Miller (Epidemic Intelligence Service Officer, Centers for Disease Control and Prevention) for manuscript review and insightful feedback. The team at Altius Institute for Biomedical Sciences: Rebecca L. Bruders, Amanda C. Gale, Clementine B.M Green, Suman Grewal, Muhammad H. Halimun, Kneshay N. Harper, Jessica M. Halow, William A. Isner, Audra K. Johnson, Jessica N. Kunder, Lauren E. Mitchell, Jemma S. Nelson, Alex S. Nguyen, Sofia E. Olsson, Sadie L. Patraw, Tobias F.C. Ragoczy, Ashly M. Senske, Julia Wald.

## Author Contributions

Conceived and designed the study: MF, HO, KA, MP, SL, IP, TB, LF

Collected or curated the data: KA, LF, SL, PR, HX, SMB, RP, ML, SE, SS, PM, ALG, LS, CF, ER, WZZ, LG, MT, JL, EM, MT, DN, DB, MH, EG, TN, JDR, JLR, JS, ET, GM, PD, DM, HG, AS, JMP, DR, LMT

Conducted the analysis: MF, MP, SL

Advised on the analysis: RB, IP, TB, KA, HO

Drafted the manuscript: MP, SL

Reviewed and edited the manuscript: All authors

## Ethics Approval

The Washington State Institutional Review Board designated this study as exempt. Sequencing and analysis of samples from the Seattle Flu Study was approved by the Institutional Review Board (IRB) at the University of Washington (protocol STUDY00006181). Sequencing of remnant clinical specimens at UW Virology Lab was approved by the University of Washington Institutional Review Board (protocol STUDY00000408).

## Data Availability

Requests to access these data are handled by the Washington Department of Health.

## Supplementary Materials

**Figure S1:**
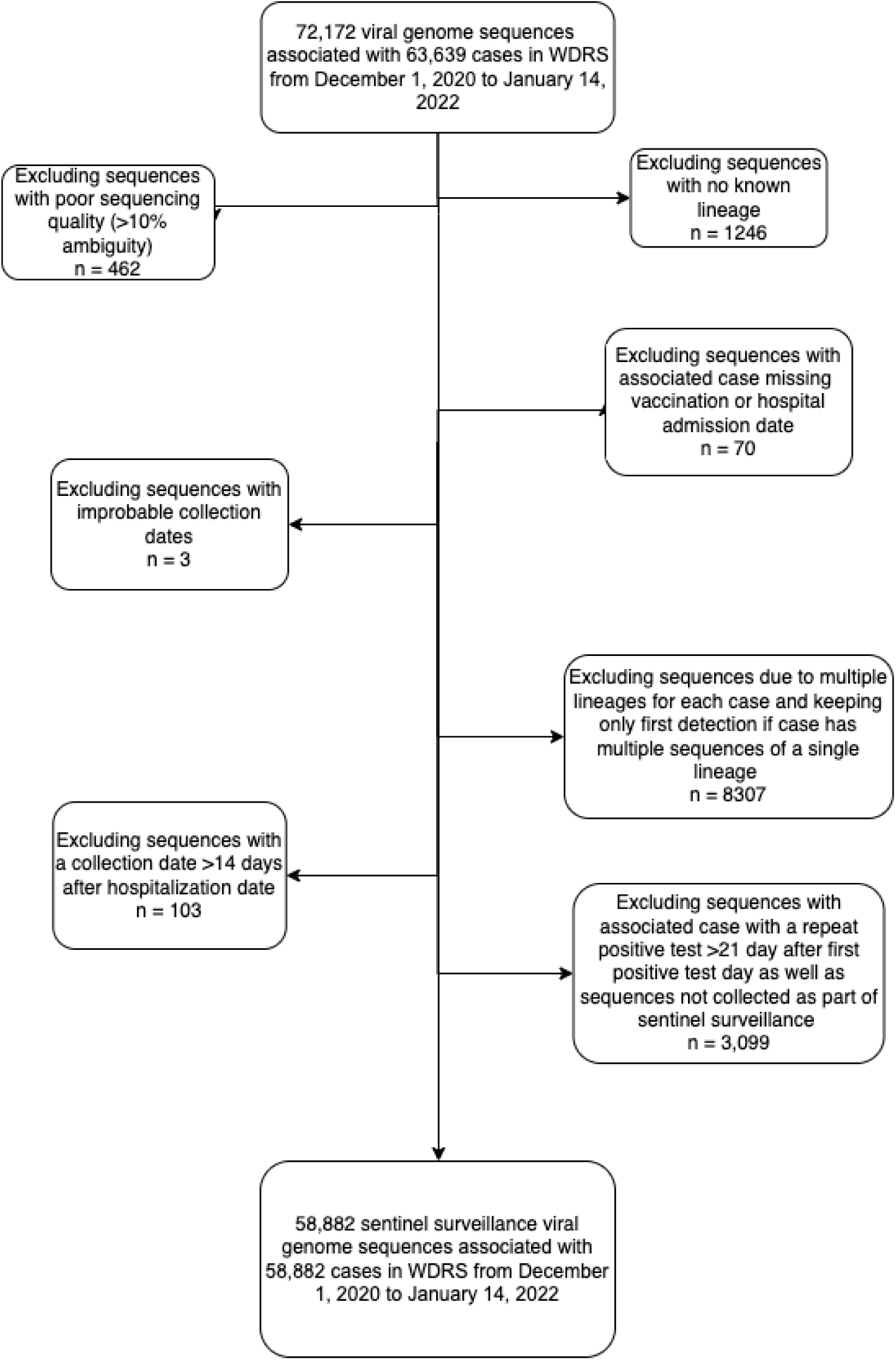
Inclusion flow diagram for study population.

**Figure S2:**
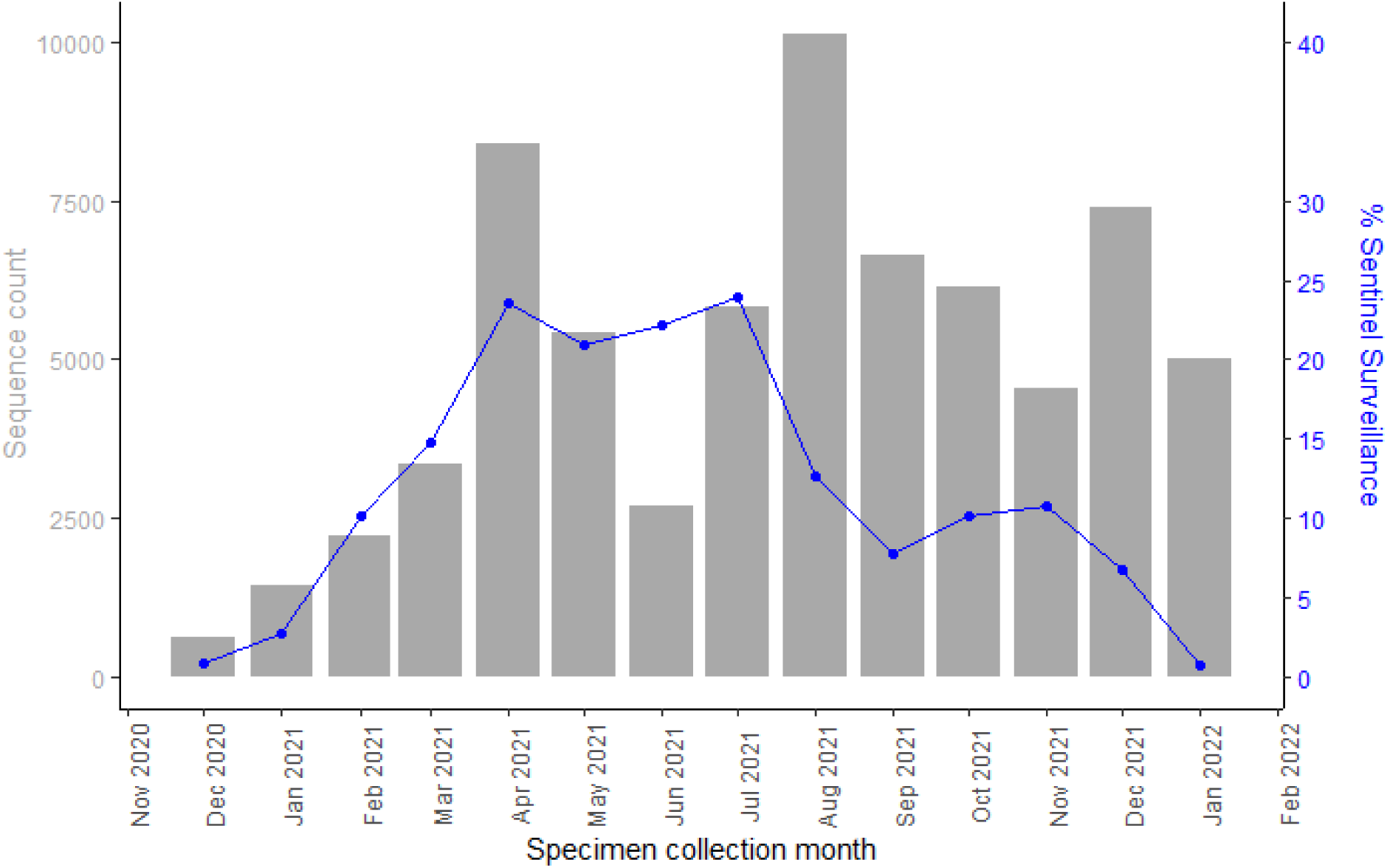
Proportion of total SARS-CoV-2 cases in Washington sequenced over time as part of sentinel surveillance. Bars represent total sequence count while blue line represents percentage of total SARS-CoV-2 positive cases in Washington that sequenced as part of sentinel surveillance for each time period. Specimens submitted from sentinel labs have decreased in January 2022 due to lab capacity during the omicron peak.

**Figure S3:**
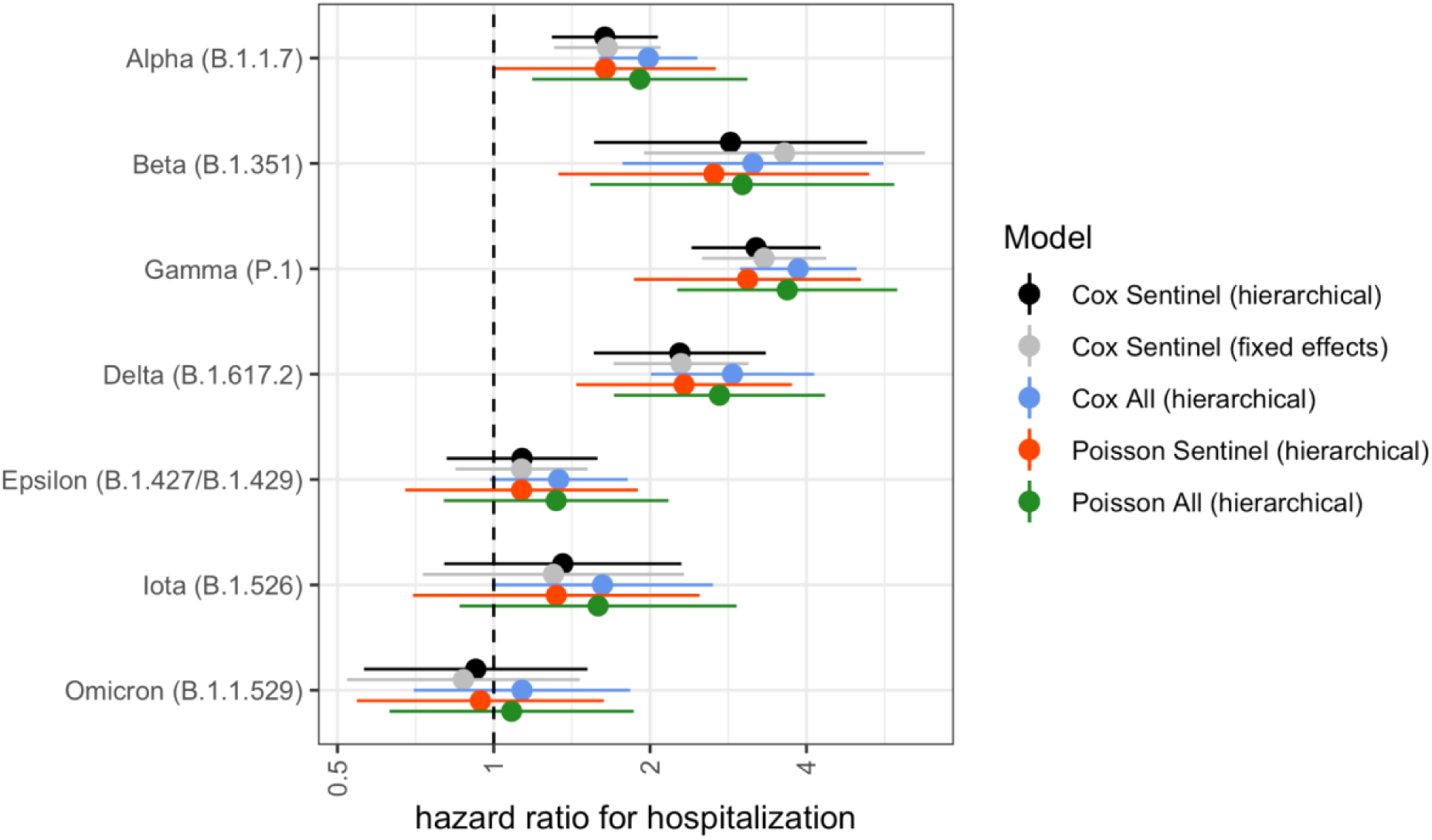
Risk of Hospitalization by Variant Lineage via differing model selection. Error bars represent 95% CI. “Hierarchical” refers to a model with mixed effects and “Sentinel” refers to the sample restricted to only to cases collected through sentinel surveillance. “Fixed effects” describes all model covariates being treated as fixed effects and “All” refers to the entire dataset from Dec 1, 2020 to Jan 14, 2022, irrespective of participation in sentinel surveillance.

**Figure S4:**
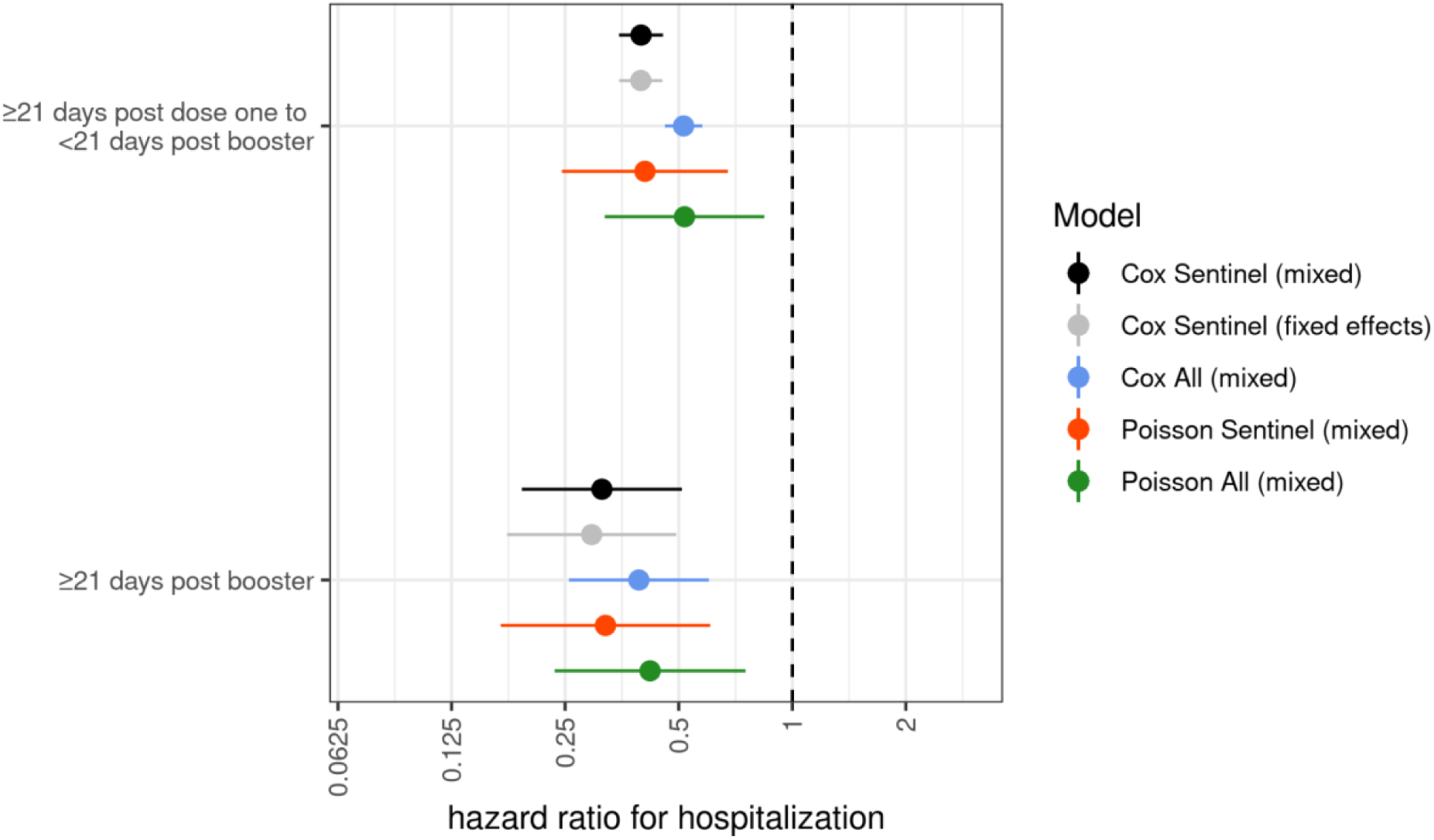
Risk of Hospitalization by vaccination dosage and technology via differing model selection. Error bars represent 95% CI. “Mixed” refers to a model with mixed effects and “Sentinel” refers to the sample restricted to only to cases collected through sentinel surveillance. “Fixed effects” describes all model covariates being treated as fixed effects and “All” refers to the entire dataset from Dec 1, 2020 to Jan 14, 2022, irrespective of participation in sentinel surveillance and includes targeted sequencing of suspected breakthrough infections.

**Table S1:** Hospitalizations by Vaccination Status.

|  | Other |  | Alpha |  | Delta |  | Gamma |  | Omicron |  |
| --- | --- | --- | --- | --- | --- | --- | --- | --- | --- | --- |
| Hospitalized | Yes | No | Yes | No | Yes | No | Yes | No | Yes | No |
| Unvaccinated to <21 days post dose one | 114 | 4975 | 224 | 8188 | 837 | 22271 | 102 | 1844 | 16 | 1984 |
| ≥21 days post dose one to <21 days post booster | – | – | 9 | 300 | 261 | 9173 | 10 | 144 | 16 | 2793 |
| ≥21 days post booster | – | – | – | – | 11 | 554 | – | – | 4 | 549 |

Categories with less than 4 hospitalizations were excluded.

**Table S2:** Adjusted Cox Proportional Hazards Estimates for Risk of Hospitalization for Omicron vs Delta.

| Characteristics | Hospitalization |  |
| --- | --- | --- |
|  | HR | 95% CI |
| *Lineage* |  |  |
| **Delta** | REF |  |
| **Omicron** | 0.34 | (0.23-0.50) |
| *Vaccination\* Lineage* |  |  |
| *Delta* |  |  |
| **Unvaccinated to <21 days post dose one** | REF |  |
| **≥21 days post dose one to <21 days post booster** | 0.40 | (0.34-46) |
| **≥21 days post booster** | 0.30 | (0.17-0.52) |
| *Omicron* |  |  |
| **Unvaccinated to <21 days post dose one** | 0.35 | (0.20-0.62) |
| **≥21 days post dose one to <21 days post booster** | 0.22 | (0.13-0.37) |
| **≥21 days post booster** | 0.18 | (0.08-0.39) |

\*Additional model covariates include: sex, age (in 10 year bins), calendar week. Each variant lineage category risk estimate uses the “Unvaccinated to <21 days post dose one” vaccination group in cases infected with Delta as the reference group.

